# Managing Cancer and Living Meaningfully Therapy Delivered as a novel remote intervention in individuals diagnosed with a Primary Central Nervous System Tumor

**DOI:** 10.64898/2026.01.07.26343618

**Authors:** Alvina Acquaye-Mallory, Gary Rodin, Madhura Managoli, Kimberly R. Robins, Macy L. Stockdill, Heather E. Leeper, Elizabeth Vera, Tito Mendoza, Amanda L. King, Michelle L. Cassidy, Mark R. Gilbert, Terri S. Armstrong

## Abstract

**Background:** Primary central nervous system (CNS) tumors affect patients’ psychological well-being and quality of life. Individualized approaches, such as Managing Cancer and Living Meaningfully (CALM), have shown potential in advanced cancers for improving these outcomes.

**Aims:** This study assessed the effects and feasibility of CALM delivered remotely to a diverse cohort of patients with a primary CNS tumor.

**Methods:** Patients completed 3-6 remote CALM sessions focusing on 4 interrelated domains. Depression, death anxiety, attachment style, and quality of life were assessed at study enrollment, 3-months, and 6-months into the intervention.

**Results:** Of the 19 patients enrolled, 15 (79% retention rate) completed the study. Most patients had a high-grade (47%) tumor, mainly diagnosed in the brain (60%). The median age was 44 years (range, 24-70). Feasibility was demonstrated through adherence to completing outcome questionnaires and a high level of patient satisfaction (100% found it worthwhile). Although no statistically significant changes were seen in depression, death anxiety, attachment anxiety, or quality of life (p > 0.05; g = −0.09 to 0.78) at any measured time, a clinically meaningful decrease in depression was observed at the 6-month point (mean difference = −3.36, p = 0.13) among spine tumor patients.

**Conclusions:** This study demonstrated that delivering CALM via telehealth is feasible, as evidenced by high compliance, low attrition, and acceptability among patients diagnosed with CNS tumors. The findings indicated meaningful reductions in depressive symptoms among patients with spinal cord tumors. These preliminary positive findings justify further evaluation of the feasibility and effectiveness of CALM in a larger sample.

**Trial registration:** ClinicalTrials.gov ID NCT04852302

## BACKGROUND

Patients diagnosed with a primary central nervous system (CNS) tumor often have shortened survival outcomes ^[1, 2]^ and significant long-term psychological challenges, leading to a reduced quality of life and shortened survival outcomes ^[1, 2]^ if these challenges remain unaddressed. Studies have shown that clinically significant depression affects over 44% of patients with primary CNS tumors ^[1]^. Despite the absence of a universally accepted intervention for depression in patients with primary CNS tumors, personalized treatments tailored to patients’ needs have proven beneficial ^[3, 4]^. However, the difficulty in distinguishing between tumor-related symptoms^[5]^ and depressive symptoms can delay appropriate treatment, potentially resulting in underutilization of psychotherapeutic interventions^[6]^.

Patients with primary CNS tumors frequently experience rapid changes in symptoms and neurocognitive difficulties ^[7]^, uncertainties and existential distress, compounded by the presence of other comorbid mental health issues ^[2]^. Addressing the emotional burden that these patients face, while simultaneously managing tumor-related symptoms that can be a barrier to psychotherapeutic compliance, is essential ^[8]^. Standard treatments for depression may limit CNS tumor patients’ participation in psychotherapeutic trials due to physical limitations, including neurologic and cognitive issues, which make frequent travel to clinical centers difficult. There may also be variability in support needed based on tumor recurrence or disease progression, which highlights the need for flexible interventions adapted to the patient’s treatment phase^[9]^. The involvement of knowledgeable staff familiar with CNS-specific challenges is vital to providing effective support^[10]^. The general guidelines for treating depression in the general population are applicable to patients with primary CNS tumors, but there may need to be tailored interventions to their unique circumstances ^[8, 9]^. Additionally, treatments can incorporate flexible modes of administering therapy, including teletherapy, which has shown benefits in reducing psychological distress, depression and enhancing the quality of life in patients with cancer ^[11, 12]^.

The CALM (Managing Cancer and Living Meaningfully) intervention is a brief semi-manualized psychotherapeutic approach that was developed to mitigate the psychological distress commonly faced by patients with advanced cancer, which can lead to the development of depression if left untreated. Over a course of 3 to 6 months, patients engage in sessions with a CALM therapist addressing 4 key domains: symptom management and communication with healthcare providers, changes in self and relationships with close others, sense of meaning and purpose, and considerations of future and mortality. One study has identified the benefits of CALM in malignant glioma patients ^[13]^, but further research is necessary to understand its effectiveness in the broader population of patients with CNS tumors across the illness trajectory. Therapists play a crucial role in helping to create a reflective space supporting mentalization, the ability to consider multiple perspectives (i.e., current realities of disease vs. impending mortality) when discussing different aspects of a patient’s life ^[14]^.

Our primary objective for this CALM trial was to evaluate its efficacy, when delivered as a remote intervention, in reducing depression among patients with primary CNS tumors. Additionally, secondary objectives explored the feasibility of remote CALM sessions and assessed its impact on death anxiety, along with exploratory aims to evaluate CALM’s influence on quality of life and interpersonal relationships over 3 and 6 months.

## METHODS

This study received approval from the National Institutes of Health Institutional Review Board (STUDY000293) in April 2021, was registered on clinicaltrials.gov (NCT04852302) and conforms to the U.S. Federal Policy for the Protection of Human Subjects. Eligibility criteria for participation included: a) adults diagnosed with a primary CNS tumor who were undergoing standard of care or experimental treatment, b) who had a life expectancy of at least 3-months from study entry that would allow them to participate in the 3 required sessions, and c) the ability to understand and sign informed consent. Patients without access to electronic devices (smartphone, computer, or tablet) for remote sessions were ineligible to participate. Patients being seen in the Neuro-Oncology Branch (NOB) clinic who reported distress were either referred by clinical teams or identified by the research team as potential candidates for eligibility screening. The research team confirmed the patient’s capacity to complete at least 3 sessions within 3 months with the clinical teams prior to consent. During the informed consent process, patients verbally indicated their interest in participating once the study was reviewed. Patients provided written informed consent to enroll in the study. Once consent was signed, patients received an email detailing their assigned CALM therapist and links to baseline measures, which needed to be completed before CALM sessions could begin.

Patient-reported outcome (PRO) measures assessed depression, quality of life, death anxiety, attachment security, and perceived benefit of clinical care, with follow-up assessments at 3- and 6-month time points. After completing the baseline questionnaires, the CALM therapist scheduled the patients’ first session. Patients completed 6 biweekly, audio-recorded remote sessions lasting approximately 45-60 minutes, which focused on 4 interrelated domains: a) Symptom management and communication with healthcare providers, b) Changes in self and relation with close others, c) Sense of meaning and purpose, d) Future and mortality.

Patients had the option to decline the recording of sessions. Additionally, to align with the CALM guidelines, booster sessions could be offered to those who wished to revisit any distressing domains. In the 1^st^ session, the CALM therapist obtained contact details for a patient-identified emergency contact and a local provider, and prior to each session, the patient’s location was verified. Given the remote nature of sessions, in instances where a patient exhibited signs of danger to themselves or others, a safety plan including coping strategies and emergency contact information was developed for the therapist’s guidance. Following the 3^rd^ session, all patients were asked about their interest in continuing to the subsequent 3 sessions, and qualitative interviews were conducted with select patients to gauge their overall experience with the CALM intervention. The study process is illustrated in *Figure 1*.

**Figure 1.**
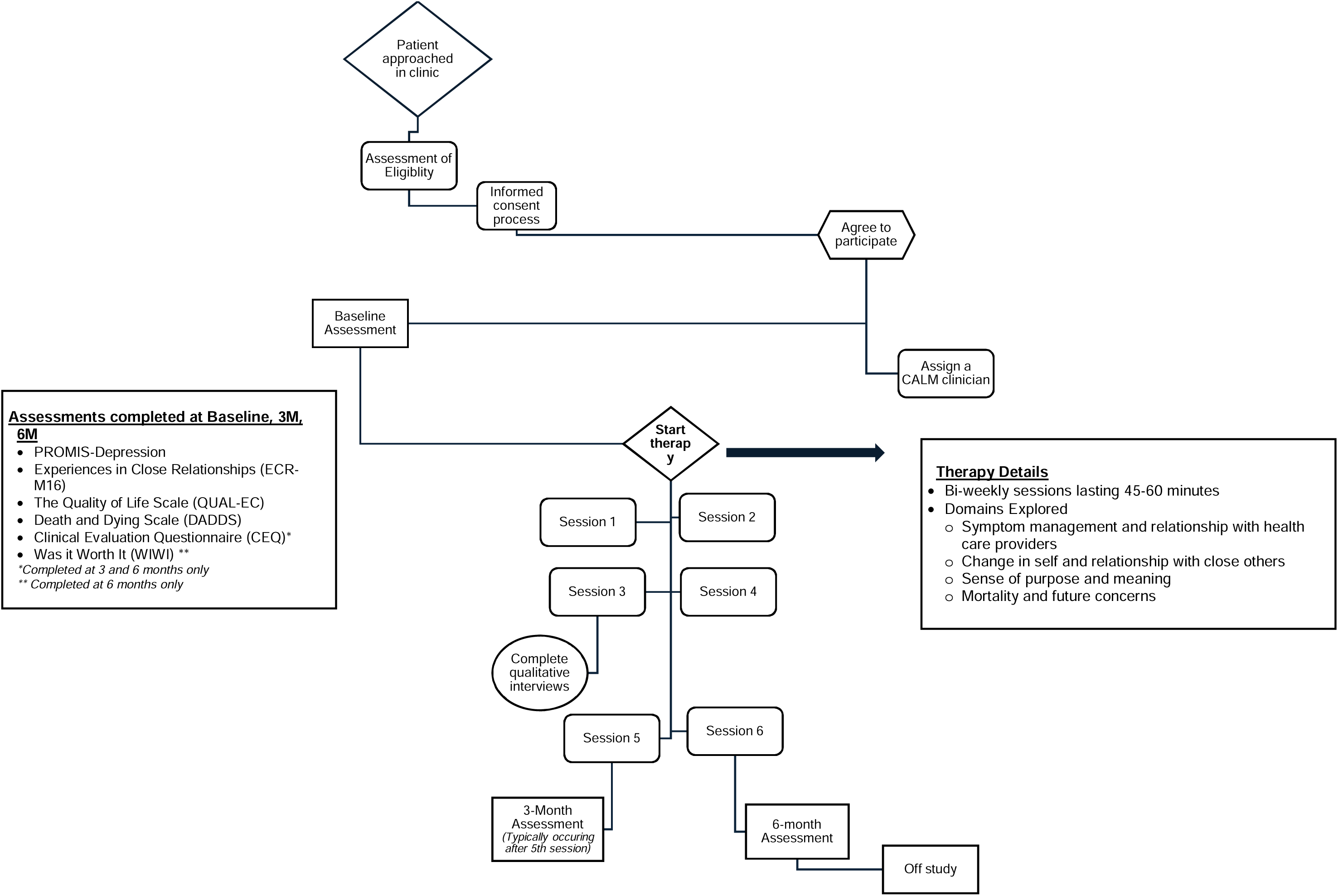
CALM Study Process

To qualify to deliver the CALM intervention, therapists (healthcare professionals involved in the care of patients with primary CNS tumors) were required to attend a CALM training workshop and complete 2 training cases (encompassing 3 to 6 sessions per patient, under supervision). AAM and HL were the 2 CALM therapists, while MS conducted the qualitative interviews.

### Measures

#### The Patient-Reported Outcomes Measurement Information System (PROMIS)-Depression

The PROMIS Depression short-form scale comprises 8 items assessing patients’ depressive symptoms within the past 7 days^[15]^. Patients report symptoms on a 5-point scale, with responses ranging from “Never” to “Always.” Raw scores are translated into *T*-scores to identify depression severity in patients. The PROMIS score thresholds are 50 (SD=10), representing the average score of the US general population; 55-59 is mild, 60-69 is moderate, and 70 and over is severe.

#### The Death and Dying Distress Scale (DADDS)

The DADDS is a 15-item instrument validated for use in advanced cancer patients. It has been shown to have two factors: Finitude, which assesses distress about the shortness of time, and Dying, which assesses distress about the process of dying and death itself ^[16, 17]^. Patients report their distress in the past 2 weeks on each item on a scale from 0 to 5, with 0 being “I was not distressed about this thought or concern” and 5 being “I experienced extreme distress.” Scores can range from 0 to 75, with higher scores reflecting higher death-related distress.

#### The Quality of Life-Cancer Scale (QUAL-EC)

The QUAL-EC is a 17-item measure that assesses the quality of life for patients nearing the end of life. It has 4 subscales (symptom control, relationship with healthcare provider, preparation for end-of-life, and life completion) and has been found to have good internal consistency ^[18]^. It has also been validated in advanced cancers. Items are added to show subscale scores, with higher scores representing a higher quality of life.

#### The modified and brief Experiences in Close Relationships (ECR-M16)

The ECR-M16 measures 16 items that focus on identifying patients’ attachment styles in close relationships with others. Patients rate items from 1 to 7, 1 being “Disagree” and 7 being “Agree,” to identify anxious or avoidant attachment styles. Items are reverse-scored and averaged, with higher scores representing attachment insecurities and lower scores representing more security in how patients perceive their close relationships. The measure has shown reliability and validity in advanced cancers^[19]^.

#### The modified version of the Clinical Evaluation Questionnaire (CEQ)

The CEQ comprises 8 items that assess the benefit of the CALM intervention. Patients completed the CEQ only at the 3 and 6-month time points by rating the perceived helpfulness of their interactions with a CALM therapist in addressing common concerns identified by cancer patients in adjusting to their diagnosis ^[20]^. Items were rated on a 5-point Likert scale, from 0 being “Not at all” to 4 being “Very much,” with scores summed to create a total score ranging from 0 to 60. Higher scores indicate more benefit of the intervention in addressing common concerns. The CEQ has shown high internal consistency in advanced cancers ^[20]^.

#### Was It Worth It (WIWI) Questionnaire

The WIWI is a 4-item questionnaire that identifies whether participation in a study was “worthwhile”. Patients responded Yes/No to each item on the WIWI, which was completed at the 6-month time point.

### Statistical Methods

Demographic and clinical characteristics were collected at study entry. For purposes of this analysis, 2 patients who enrolled in CALM had both brain and spinal cord tumors, but were included in the spine tumor category for analysis as they both initially presented with tumors in the spine. Descriptive statistics were used to summarize the patient sample and provide PRO summary scores at baseline, 3-months, and 6-months of participating in the CALM intervention. Paired-sample *t*-tests and paired-sample proportion tests (*z*-tests) were used to compare the PROs from baseline to 6 months, baseline to 3 months, and 3 months to 6 months, with effect size reported using Hedges’ g and Cohen *h*, respectively. All statistical analyses were performed utilizing IBM SPSS Version 29.0.1.1 and statistical tests were two-sided with a significance level of 0.05 ^[21]^.

## RESULTS

The demographic and clinical characteristics are provided in Table 1. A total of 19 patients enrolled in the study; 2 patients withdrew (1= busy schedule, 2= not ready to start any form of therapy), and only 15 patients completed all requirements for analysis. The sample included slightly more males (53%) who identified as White (73%), with a median age of 44 years (minimum, maximum: 24,70 years). A diagnosis of high-grade tumors (47%) was common, and 60% of the tumors were in the brain. Prior to starting CALM, 33% had experienced tumor recurrence, and at baseline, 93% of patients were not receiving active treatment.

**Table 1.**
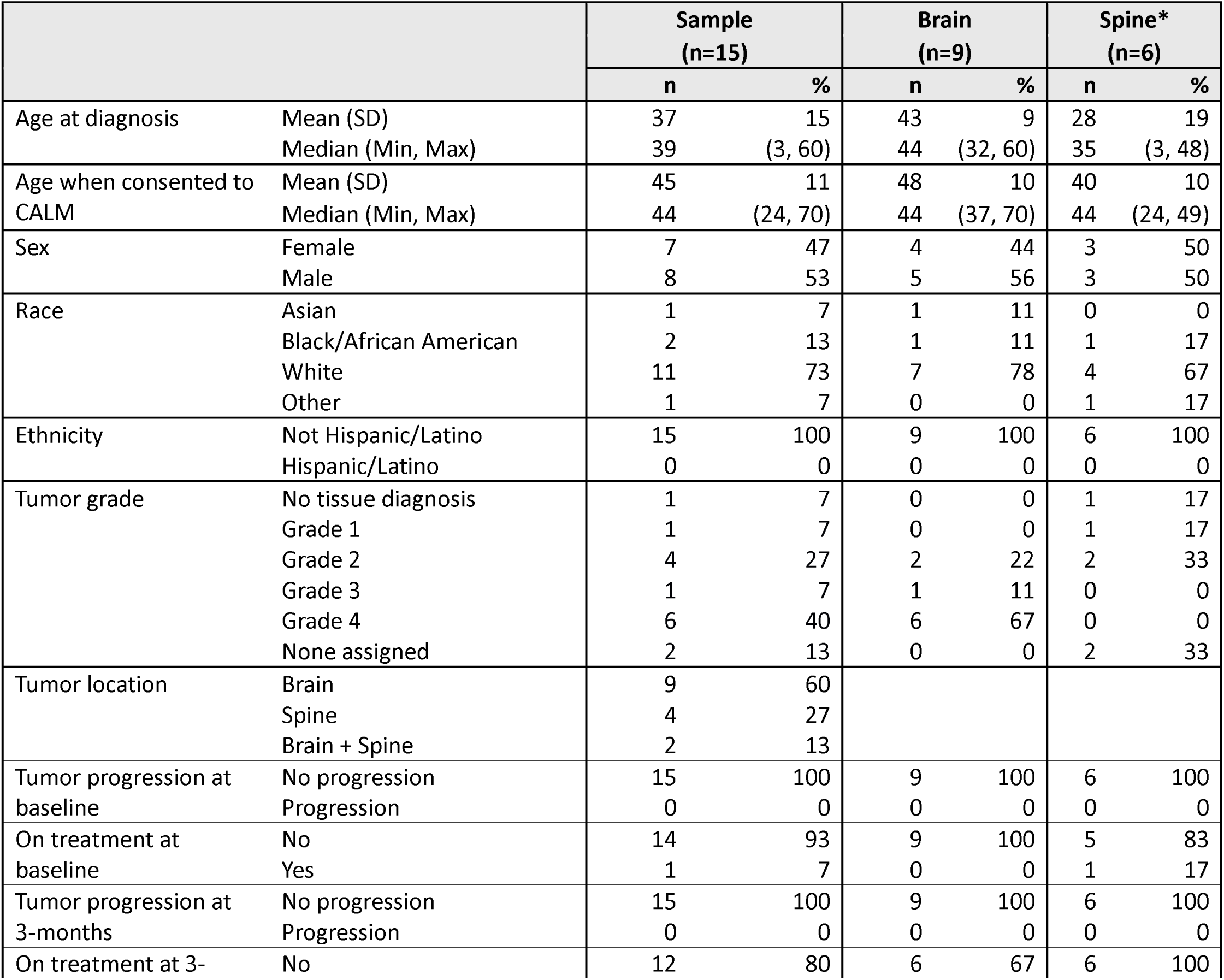
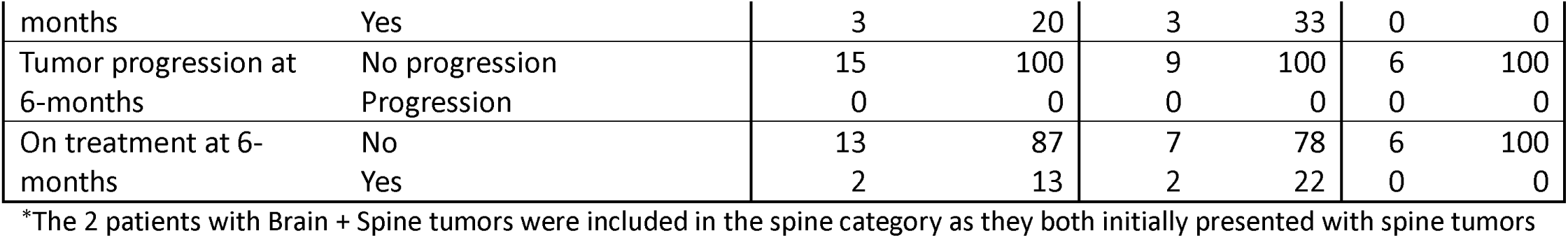
Demographic and Clinical Characteristics.

### PROMIS-Depression

Table 2 displays the PROMIS-Depression scores for the total sample. The sample reported slightly higher scores (Mean [M]=54.79, standard deviation [SD]=8.78) than the general population, indicating mild depressive symptoms at baseline. Moderate to severe depressive symptoms were experienced by 27%. Although no significant changes were observed, a steady decline is evident from baseline to 6 months, with scores dropping by −1.81 points (95% CI: −4.47, 0.86). Patients with spine tumors demonstrated a more substantial drop, with a mean difference of −3.36 points (95% CI: −8.09, 1.37), compared to brain tumor patients (mean difference = −0.77 (95% CI: −4.60, 3.06)) from baseline to 6 months.

**Table 2.**
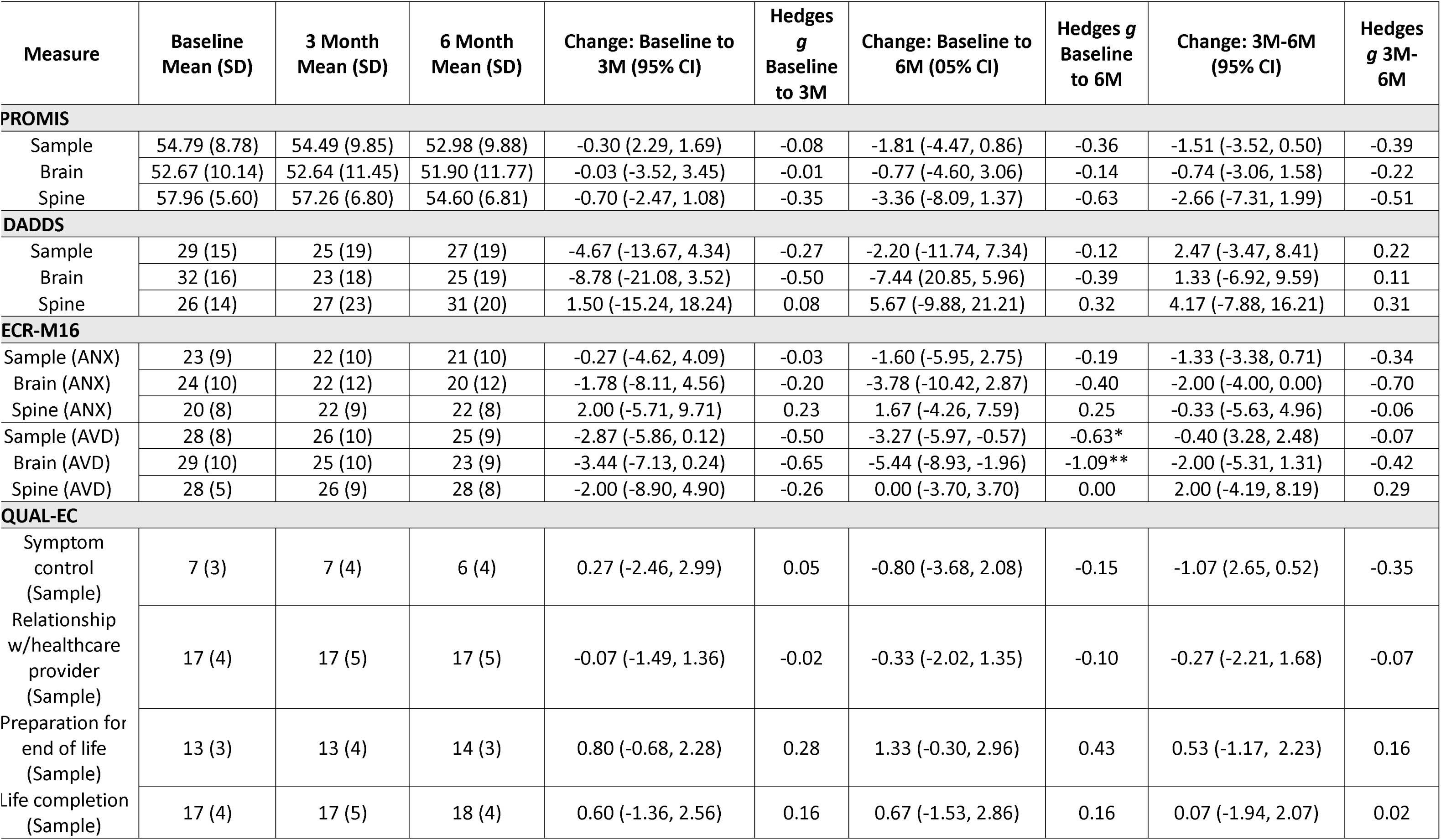
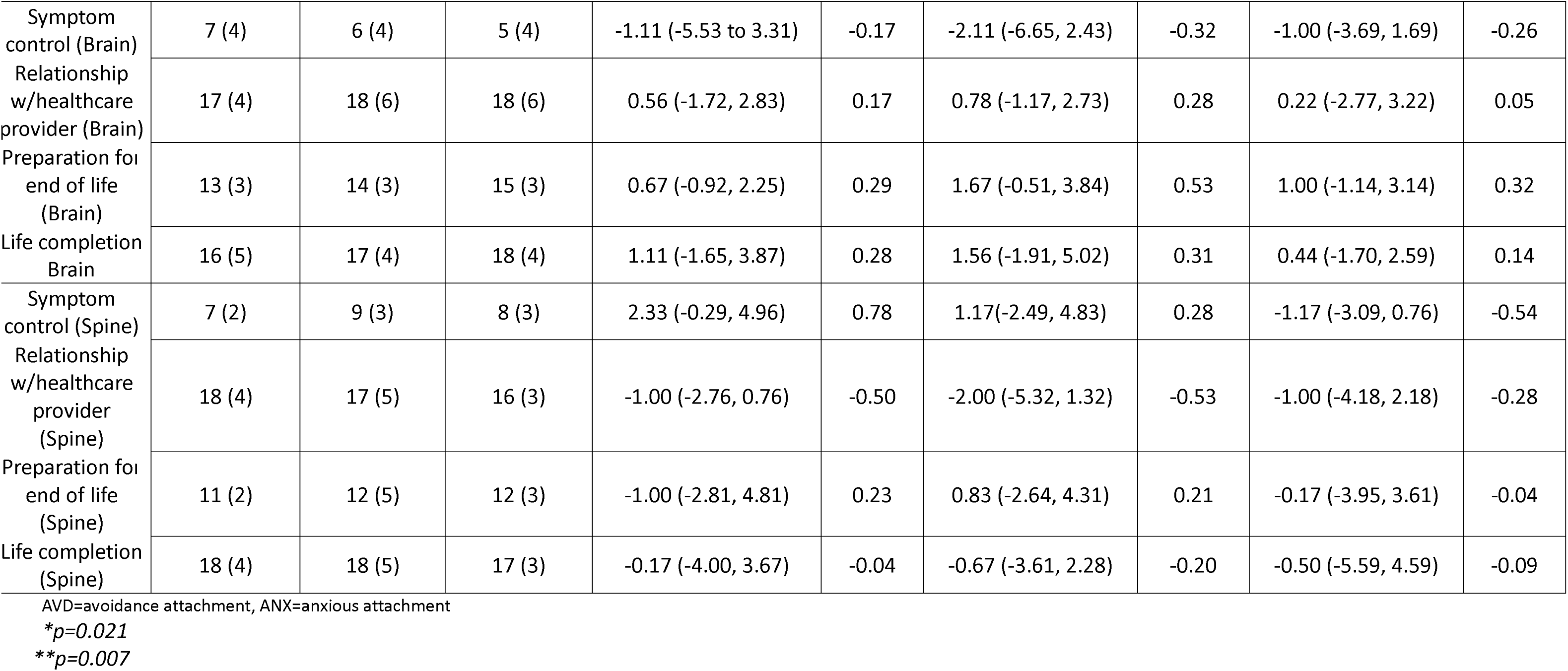
Mean Changes in PROs.

### Secondary and Exploratory Findings

#### Feasibility of CALM

The study demonstrated feasibility by showing that patients were able to complete the CALM intervention remotely with 100% compliance on PRO questionnaires both at baseline and 3 months. At 6 months, 5 out of 6 PROs were completed, as one individual did not complete the WIWI (Table 3). All patients consented to sessions 4 to 6. Patients reported positive outcomes on the WIWI questionnaire at 6 months. Eighty percent indicated they would choose to participate in the CALM intervention again. Additionally, 73% noted a change in their quality of life due to their participation in the CALM intervention, while all patients found it to be ‘worthwhile’ and would ‘recommend’ it to others experiencing similar health challenges.

**Table 3.** WIWI 6-month Descriptives.

|  |  | Total Sample<br>(n=14) |  | Brain Tumor<br>(n=8) |  | Spine Tumor<br>(n=6) |  |
| --- | --- | --- | --- | --- | --- | --- | --- |
|  |  | n | % | n | % | n | % |
| If you had to do it over, would you choose to participate in the CALM intervention again | No | 3 | 20 | 1 | 13 | 2 | 33 |
|  | Yes | 11 | 80 | 7 | 88 | 4 | 67 |
| Overall, did your quality of life change by participating in the CALM intervention? | No | 4 | 27 | 2 | 25 | 2 | 33 |
|  | Yes | 10 | 73 | 6 | 75 | 4 | 67 |
| Was it worthwhile for you to participate in the CALM intervention? | No | 0 | 0 | 0 | 0 | 0 | 0 |
|  | Yes | 14 | 100 | 8 | 100 | 6 | 100 |
| Would you recommend the CALM intervention to others with your illness? | No | 0 | 0 | 0 | 0 | 0 | 0 |
|  | Yes | 14 | 100 | 8 | 100 | 6 | 100 |

#### The Death and Dying Distress Scale (DADDS)

When examining death anxiety in patients (shown in Table 2), the DADDS scores showed that the sample experienced moderate levels of death anxiety at baseline (M=29, SD=15), with brain tumor patients (M=32, SD=16) reporting higher distress scores related to death and dying compared to spine tumor patients (M=26, SD=14). Overall, the DADDs remained stable over the 6-month follow-up period, showing no significant improvements or declines in death anxiety; the largest change in scores occurred from baseline to 3 months in the sample, with a mean difference of −4.67 (95% CI: −13.67,4.34). For brain tumor patients, findings show moderate reductions in the DADDs over 6 months (*g*=-0.39, −0.50). Conversely, spine tumor patients exhibited increased DADDs scores between baseline, 3 months, and 6 months.

#### The modified and brief Experiences in Close Relationships (ECR-M16)

The ECR-M16 findings can be found in Table 2. More patients reported having avoidant attachment (M=28, SD=8) in close relationships compared to anxious attachment (M=23, SD=9). A significant decrease in avoidant attachment was observed in the sample from baseline to 6 months, with a mean difference of −3.27 (95% CI: −5.97, −0.57). Additionally, patients with brain tumors exhibited a significant mean difference of −5.44 (95% CI: −8.93, −1.96), indicating a reduction in avoidant attachment over this period. Conversely, spine tumor patients did not demonstrate significant changes in avoidant attachment from baseline to 6 months. Although no significant changes were noted in the anxious attachment style, the overall scores for the sample showed a steady decline from baseline to 6 months, yielding a mean difference of −1.60 (95% CI: −5.95, 2.75). Furthermore, patients with brain tumors experienced a slight decrease in anxious attachment scores, with a mean difference of −3.78 (95% CI: −10.42, 2.87). In contrast, those with spine tumors showed an increase in anxious attachment scores with a mean difference of 1.67 (95% CI: −4.26, 7.59) from baseline to 6 months.

#### The Quality of Life-Cancer Scale (QUAL-EC)

Table 2 presents the total sample and the breakdown of the QUAL-EC subscales by tumor location. Across the sample and among both brain and spine tumor patients, no statistically significant differences were observed in symptom control, relationships with healthcare providers, preparation for end-of-life, or life completion scores over time. The effect sizes for the total sample were consistently small. Patients had higher scores (M=17, SD=4) on the 2 subscales (relationships with healthcare providers and life completion). Symptom control was rated as the lowest domain affecting patients’ quality of life at all 3 time points. Although spine tumor patients showed a trend towards improved symptom control at 3 months (2.33, 95% CI: −0.29, 4.96), this did not reach significance.

#### The modified version of the Clinical Evaluation Questionnaire (CEQ)

Regarding the CEQ results at 3 months, the most frequently reported benefits of CALM included ‘talking and feeling understood about how cancer has affected my life’ and being able to ‘better express and manage my feelings,’ with 53% reporting it helped ‘quite a bit.’ Patients also highlighted being able to ‘express and manage my fears about dying’ at 46%. At the 6-month mark, patients continued to report improved expression and management of feelings, followed by being able to ‘talk about what will happen to my family’ at 50% and being able to ‘deal with changes in my relationship as a result of cancer’ at 47%. Descriptives for the CEQ is present in Table 4.

**Table 4.** CEQ 3-month and 6-month Descriptives.

|  |  | Total Sample<br>(n=15) |  | Brain Tumor<br>(n=9) |  | Spine Tumor<br>(n=6) |  |
| --- | --- | --- | --- | --- | --- | --- | --- |
| **3-mo.** | Mean (SD) | 34 | 14 | 37 | 12 | 30 | 17 |
|  | Median (Min, Max) | 36 | (10,54) | 36 | (22,54) | 30 | (10,50) |
| **6-mo.** | Mean (SD) | 33 | 14 | 35 | 15 | 31 | 14 |
|  | Median (Min, Max) | 32 | (9,54) | 32 | (9,54) | 34 | (9,45) |

## DISCUSSION

This study is 1 of only 2 reported studies that utilized a remote CALM intervention for patients with primary CNS tumors ^[13]^. Our findings indicated high satisfaction and compliance with the sessions and low attrition, demonstrating the feasibility of administering CALM remotely to patients with primary CNS tumors. Although the hypothesized effectiveness of CALM in reducing depression among patients with a primary CNS tumor did not achieve statistical significance, a slight improvement in depressive scores over time was observed, with significant differences noted among spine tumor patients. Additionally, exploratory findings revealed significant reductions in avoidant attachment styles among the overall sample and among patients with brain tumors, suggesting a shift toward more secure attachment in personal relationships. These results, combined with the paucity of existing literature on the application of CALM in patients with a primary CNS tumor, underscore the need for further exploration of the intervention’s impact and potential benefits for this unique patient population.

Additional research is needed to establish the efficacy of CALM in this population, although our findings align with other CALM studies, which also found improvements in psychological well-being and depressive symptoms ^[13, 22]^. Interestingly, despite a smaller sample size of only 15 patients, our effect size was stronger than anticipated, highlighting what further insights might emerge with increased enrollment toward our original goal. The exploratory data are consistent with the hypothesized view of CALM’s beneficial impact on psychological outcomes. Patients with primary CNS tumors face distinct psychosocial challenges influenced by tumor location and social factors that play a pivotal role in their coping abilities^[23, 24]^. It is worth noting that many patients did not perceive the intervention as applicable during screening, reflecting a common issue in psychotherapeutic trials where patients’ uncertainty about treatment benefits may hinder participation^[25]^.

### Study Limitations

This study had limitations, notably the small sample size. Low accrual was impacted by the study being initiated during COVID and the limited clinic availability, which impacted the number of patients that could be seen. Future research should prioritize enrolling an adequate number of patients to demonstrate feasibility and to delineate the psychological benefits of CALM, particularly for patients with cancer types who might be excluded due to tumor-related challenges. Additionally, while convenient, the virtual components of the intervention may have hindered rapport building that is essential for effective therapy. It is possible that a blended approach incorporating telehealth and in-person options could enhance patient experience and outcomes. Given that most patients were not receiving treatment at baseline (entry into the study), future studies should aim to enroll a diverse sample encompassing various treatment phases. The needs of patients at the end of life likely differ from those with stable disease or undergoing treatment, so exploring these distinctions could provide valuable insights into how a therapeutic intervention is tailored.

### Clinical Implications

Our findings provide valuable insight for optimizing the CALM intervention to benefit patients with a primary CNS tumor. The changes identified in patient-reported outcomes, particularly in attachment styles, emphasized the need for additional studies focusing on psychotherapeutic interventions for this often-excluded cohort. As psychological challenges evolve throughout the illness trajectory, it may be beneficial to explore tailored approaches at different phases of the disease to standardize or personalize interventions in future trials. Expanding upon the domains of CALM or proposing multi-faceted alternate treatment modalities to address patient-specific needs may yield important advancements in care.

## CONCLUSIONS

In conclusion, this preliminary study aimed to investigate the effects and feasibility of the CALM intervention to reduce depression and to improve psychological well-being in a novel group (primary CNS tumors). Contrary to our hypothesis, the results did not reveal statistically significant relationships; however, exploratory outcomes suggested significance in reducing patients’ avoidant attachment styles. Small to moderate effects on depression, anxious attachment, quality of life, and death anxiety were identified, but future studies should target a larger sample size to identify meaningful differences. CALM provides a unique experience for patients in addressing existential concerns while offering a supportive space, and with over 70% of patients reporting a change in quality of life, it is essential to address specific issues related to primary CNS tumors, particularly concerning tumor location and treatment phase.

## Data Availability

All data produced in the present work are contained in the manuscript

## AUTHOR CONTRIBUTIONS

Conceptualization: TSA, AAM, ALK, HEL, GR, EV

Data curation: EV, KRR

Formal analysis: EV, KRR, TM

Funding acquisition: TSA

Investigation: AAM, MRG, TSA

Methodology: AAM, KRR, EV, TSA, HEL, ALK

Project administration: TSA Resources: TSA, MRG

Supervision: TSA, MRG

Validation: KRR, EV, TSA, TM

Visualization: AAM, KRR

Writing – original draft: AAM

Writing – review & editing: AAM, GR, MM, KRR, MLS, HEL, EV, TM, ALK, MLC, MRG, TSA

## ACKNOWLEDGEMENTS

We thank the patients and their families for their time and effort in making this work possible. This research was supported in part by the Intramural Research Program of the National Institutes of Health, National Cancer Institute. Additionally, Macy Stockdill, PhD, was a post-doctoral fellow supported by the National Institute of Health, National Cancer Institute’s Intramural Continuing Umbrella for Research Experiences (iCURE) program. This research was supported [in part] by the Intramural Research Program of the National Institutes of Health (NIH). The contributions of the NIH author(s) were made as part of their official duties as NIH federal employees, are in compliance with agency policy requirements, and are considered Works of the United States Government. However, the findings and conclusions presented in this paper are those of the author(s) and do not necessarily reflect the views of the NIH or the U.S. Department of Health and Human Services.

## ETHICS STATEMENT

This study conforms to the U.S. Federal Policy for the Protection of Human Subjects. Approval was received from the National Institutes of Health Institutional Review Board (STUDY000293).

## CONSENT

All patients have given informed consent.

